# A Systematic Review of Representation in Clinical Trials for Transthyretin Associated Cardiac Amyloidosis: Where Demographics Diverge from Disease

**DOI:** 10.64898/2026.02.04.26345614

**Authors:** Dimitri Ford, Akshay Chandora, Chima Amadi, Niral Thaker, Ridwan Azees, Matthew E. Gold, Nicolas Bakinde, Anekwe E. Onwuanyi, Marshaleen Henriques King

## Abstract

**Background:** Underrepresentation of minority groups in clinical trials worsens health disparities and reduces generalizability of results. Transthyretin-associated cardiac amyloidosis (ATTR), is a condition that disproportionately impacts some racial and ethnic groups yet the extent to which trial enrollment reflects disease burden remains unclear. Misalignment between disease prevalence and trial representation may delay treatment development and increase the economic burden of late diagnosis and mismanagement.

**Methods:** This was a systematic review of US-based ATTR clinical trials found on clinicaltrials.gov up to 2025 (search date February 12, 2025). Only completed trials with publicly available results were included. Demographic data were extracted at the trial level. An enrollment fraction (where EF = observed enrollment ÷ expected enrollment based on Cardiac Amyloidosis Registry Study (CARS) prevalence; adequacy defined EF ≥ 0.75) was calculated for each group.

**Results:** Of the 264 clinical trials on ATTR identified, 16 met inclusion criteria. African Americans/Individuals of African descent had EFs below the adequate ratio of 0.75 in all phases of the trials reviewed compared to their Asian or White counterparts. Despite the FDA Final Rule in 2017, our study showed that there was increased study demographic reporting (60% to 85.7%), but a paradoxical decline for Black participants (EF 0.29 to 0.12, p < 0.001) and other minority participants.

**Conclusions:** Black individuals remain substantially underrepresented in U.S. ATTR-CM clinical trials despite improved demographic reporting after the 2017 Final Rule. Actionable strategies, community engagement, trial-site diversification, enrollment targets, and sponsor accountability are needed to improve representativeness and expand access.

## Introduction

Transthyretin-associated cardiac amyloidosis (ATTR-CM) is a progressive and life-threatening condition. It is caused by the misfolding of transthyretin (TTR) proteins, which aggregate as amyloid fibrils in cardiac tissue and infiltrate the myocardium, leading to restrictive cardiomyopathy, heart failure, and arrhythmias [1]. ATTR-CM exists in two main forms: hereditary (variant, ATTRv) and wild-type (ATTRwt), the latter typically affecting older adults without known genetic mutations [2]. Median survival following diagnosis ranges from 2 to 6 years, with worse outcomes in those presenting with advanced heart failure. ATTRwt may account for up to 10–15% of heart failure cases with preserved ejection fraction (HFpEF) in older adults, and autopsy studies suggest a prevalence as high as 25% in men over 80 years [3]. Yet, the disease remains underdiagnosed, often mistaken for other forms of cardiomyopathy. This contributes to an average diagnostic delay of 34 months, with worse cardiac function associated with longer delays in diagnosis [4].

Clinical trials play a transformative role in reshaping the care of patients with ATTR-CM. Landmark studies, such as the ATTR-ACT trial, demonstrated that therapies like tafamidis significantly reduce mortality and cardiovascular-related hospitalizations [5]. These advances underscore the critical importance of clinical research in altering the natural history of this once-fatal disease. However, significant challenges persist. Despite advancements in diagnostics and therapeutics, disparities in outcomes remain. Evidence indicates that certain populations, particularly racial and ethnic minorities and lower socioeconomic groups, experience delayed diagnosis, reduced access to advanced therapies, and worse prognosis [6,7]. Although access to clinical trials should be equitable, multiple studies have shown underrepresentation of minority groups. In response to concerns about transparency and equity in research, the U.S. government implemented the Final Rule for Clinical Trials Registration and Results Information Submission (CTRRIS) in 2017. This policy mandates the timely registration and reporting of clinical trial data on clinicaltrials.gov [8]. While this was a step forward in promoting accountability and inclusivity, we hypothesized that a discrepancy still exists between the demographics of patients enrolled in ATTR-CM trials and those most burdened by the disease. Addressing this gap is crucial for ensuring equitable benefit from scientific advances.

## Methods

This was a systematic review using a search on clinicaltrials.gov on 12 February 2025. The terms “cardiac amyloidosis” and “amyloidosis cardiac” were used to identify the initial studies, and were then filtered to completed, U.S.-based adult studies. No time constraints were utilized. We extracted trial-level data (number enrolled, phase, year, and reported race/ethnicity and sex distributions). Trials were included if they enrolled predominantly ATTR-CM patients and publicly reported demographic results. Trials with pediatric populations or lacking demographic data were excluded.

To quantify representation, the enrollment fraction (EF) was calculated for White and Black participants only, as incidence data for other demographic groups were unavailable. EF, originally described by Murthy et al, is defined as the number of clinical trial enrollees divided by the estimated number of U.S. cases within each race, ethnicity, and gender subgroup [9].

The formula for calculating EF is as follows:

EF = Observed enrollment/ Expected Enrollment

Expected Enrollment = total number of participants enrolled x CARS observed demographic percentage.

Estimates of ATTR prevalence were obtained from the Cardiac Amyloidosis Registry Study (CARS), a multicenter registry established in 2019 that includes patients with transthyretin-related (both wild-type and variant) and light chain (AL) cardiac amyloidosis evaluated at major amyloidosis centers between 1997 and 2023 [10]. Per the CARS observational study, the prevalence of ATTR is as follows: Whites 73%, Black/African American 24%, Asian 2% other 1% and Hispanic 3%. Prevalence per sex was as follows: Men 87% and women 13%.

An EF threshold of 0.75 was selected to define adequate representation. This cutoff has been used in prior literature evaluating demographic proportionality in U.S. clinical trials to indicate near-parity with disease prevalence while allowing for expected sampling variability and logistical constraints in recruitment. An EF of 1.0 reflects perfect proportionality between trial enrollment and disease prevalence within a demographic subgroup. However, given inherent fluctuations in registry-based prevalence estimates, trial eligibility criteria, and geographic concentration of specialized centers, a tolerance range of approximately 25% below parity (i.e., EF ≥ 0.75) is considered methodologically reasonable to represent adequate inclusion. Values below this threshold indicate substantial underrepresentation relative to the disease population, while values exceeding 1.25 denote relative overrepresentation. This convention provides a pragmatic benchmark for assessing equity in trial enrollment and has been adopted in recent diversity analyses of cardiovascular and oncologic clinical research [9,11].

Descriptive statistics were generated using Python, and chi-square tests were performed to assess differences in EF across groups. A p-value < 0.05 was considered statistically significant.

## Results

This study identified 264 clinical trials on amyloidosis, of which 16 met the inclusion criteria specifically for ATTR. As shown in **Table 1**, these trials enrolled a total of 11,447 participants, with a male-to-female ratio of approximately 3:1. Among the 16 trials, five were phase 3, four were observational, four were phase 2, two were phase 4, one was a combined phase 1 and 2, and one was an interventional study without a reported phase. Of these, 10 trials were conducted before the implementation of CTRRIS in 2017, and 6 clinical trials were conducted afterward. Prior to 2017, 60% of trials reported racial demographics, while 85.7% reported racial demographics afterwards.

**Table 1.** Comparing participant demographics.

| Name | Year | Study Design | Enrollment | Male | Female | Black | White | Asian |
| --- | --- | --- | --- | --- | --- | --- | --- | --- |
| **Transthyretin Amyloidosis Outcome Survey (THAOS)** | **2008** | Observational | 6718 | 4304 | 2414 | 318 | 3441 | 295 |
| **The Effects of Fx-1006A on Transthyretin Stabilization and Clinical Outcome Measures in Patients With Non-V30M Transthyretin Amyloidosis** | **2008** | Phase 2 | 21 | 13 | 8 | Did not report | Did not report | Did not report |
| **The Effects Of Fx-1006A On Transthyretin Stabilization And Clinical Outcome Measures In Patients With V122I Or Wild-Type TTR Amyloid Cardiomyopathy** | **2008** | Phase 2 | 35 | 32 | 3 | Did not report | Did not report | Did not report |
| **Safety And Efficacy Evaluation Of Fx-1006a In Patients With V122i Or Wild-Type** | **2009** | Phase 3 | 31 | 28 | 3 | 4 | 27 | 0 |
| **Transthyretin (TTR) Amyloid Cardiomyopathy** |  |  |  |  |  |  |  |  |
| **Burden of Disease Study In Patients With Transthyretin Familial Amyloidosis Polyneuropathy (TTR-FAP) of Transthyretin Cardiomyopathy (TTR-CM) And Caregivers** | **2012** | Observational | 92 | 57 | 35 | Did not report | Did not report | Did not report |
| **Doxycycline and TUDCA in Patients With Transthyretin Amyloid Cardiomyopathy** | **2013** | Phase 1 and 2 | 40 | 29 | 1 | Did not report | Did not report | Did not report |
| **Safety and Efficacy of Tafamidis in Patients With Transthyretin Cardiomyopathy (ATTR-ACT)** | **2013** | Phase 3 | 441 | 398 | 43 | 63 | 357 | 18 |
| **A Extension Study to Evaluate Revusiran (ALN-TTRSC) in Patients With Transthyretin (TTR) Cardiac Amyloidosis** | **2014** | Phase 2 | 25 | 23 | 2 | 4 | 21 | 0 |
| **ENDEAVOUR: Phase 3 Multicenter Study of Revusiran (ALN-TTRSC) in Patients With Transthyretin (TTR) Mediated Familial Amyloidotic Cardiomyopathy (FAC)** | **2014** | Phase 3 | 206 | 158 | 48 | 104 | 95 | 0 |
| **Long-term Safety of Tafamidis in Subjects With Transthyretin Cardiomyopathy** | **2016** | Phase 3 | 1733 | 1542 | 186 | 159 | 1468 | 80 |
| **Study of AG10 in Amyloid Cardiomyopathy** | **2018** | Phase 2 | 49 | 45 | 4 | 10 | 35 | 1 |
| **Prevalence and Characteristics of Transthyretin Amyloidosis in Patients With Left Ventricular** | **2018** | Observational | 812 | 533 | 233 | 21 | 712 | 27 |
| **Hypertrophy of Unknown Etiology (TTRACK)** |  |  |  |  |  |  |  |  |
| **Efficacy and Safety of AG10 in Subjects With Transthyretin Amyloid Cardiomyopathy** | **2019** | phase 3 | 632 | 570 | 62 | 30 | 555 | 13 |
| **Long-term Monitoring of Patients With Cardiac Amyloidosis With Implantable Event Monitors** | **2020** | Not reported | 24 | 24 | 0 | 0 | 24 | 0 |
| **SGLT2 Inhibitors in Transthyretin Amyloid Cardiomyopathy** | **2022** | Phase 4 | 15 | 13 | 2 | 2 | 13 | 0 |
| **A Retrospective Validation Study To Identify Chart-Based Clinical Diagnosis Of Wild-Type Transthyretin Amyloid Cardiomyopathy (Attrwt-CM) And Non-Amyloid Heart Failure Among Patients With Heart Failure (HF).** | **2023** | Observational | 558 | 558 | 0 | Did not report | Did not report | Did not report |
| **Total** |  |  | 11371 | 8327 | 3044 | 715 | 6748 | 434 |

The overall racial demographic of enrolled participants was as follows; Whites 59.4%, Black or African Americans at 6.3%, Asians (3.8%), Latino individuals (1.9%), Native Hawaiian or Other Pacific Islanders (<0.1%), and Others (1.2%), as shown in **Figure 1**. There were 27.4% of trials that did not report racial demographic information.

**Figure 1.**
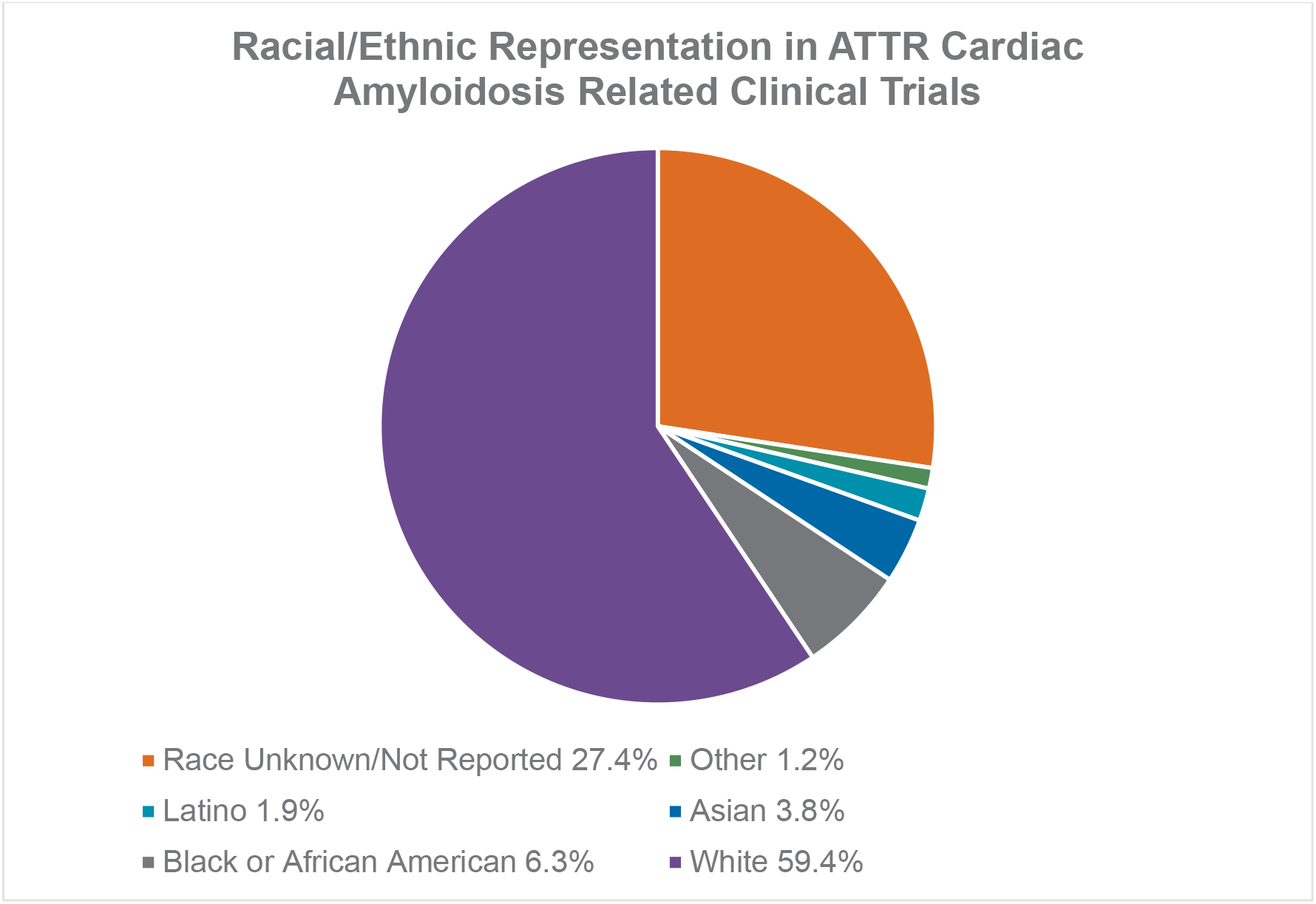
Race and Ethnicity Representation in Transthyretin Associated Cardiac Amyloidosis Trials: This chart displays the breakdown of the reported race and ethnicity of patients in clinical trials

**Table 2** shows the EF for each race to be White 0.81 (95% Confidence Interval (CI) 0.80 – 0.82), Black/ African American 0.26 (95% CI 0.24 – 0.28), Asian 1.9 (95% CI 1.73 – 2.08), (P value <0.001) and Men 0.84 (95% CI 0.83 – 0.85) and Women 2.05 (95% CI 1.99 – 2.12), (P Value <0.001).

**Table 2.** Overall Cardiac Amyloidosis Enrollment Factor Ratio Comparison Between Races.

| Group | Observed | Expected | EF ratio | 95% CI |
| --- | --- | --- | --- | --- |
| White | 6748 | 8301 | 0.813 | 0.798 – 0.822 |
| Black | 715 | 2729 | 0.262 | 0.242 – 0.278 |
| Asian | 434 | 227 | 1.912 | 1.725 – 2.075 |
| chi-square goodness-of-fit tests: p = <0.001 (0.000) |  |  |  |  |
| men | 8327 | 9893 | 0.842 | 0.831 – 0.849 |
| women | 3044 | 1478 | 2.060 | 1.985 – 2.115 |
| chi-square goodness-of-fit tests: p = <0.001 (0.000) |  |  |  |  |

**Table 3a.**
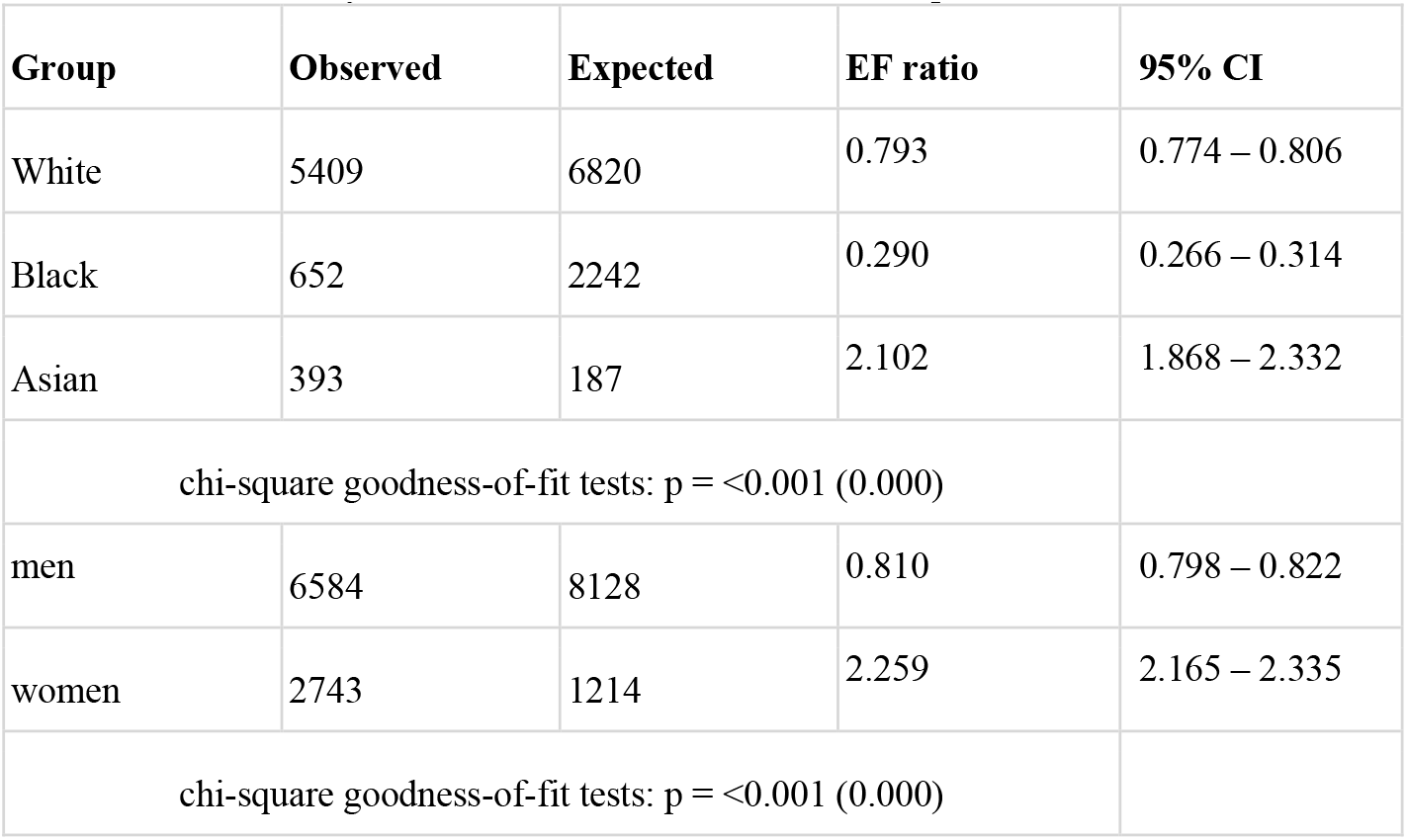
Cardiac Amyloidosis Enrollment Factor Ratio Comparison Between Races Prior to 2017.

| **Group** | **Observed** | **Expected** | **EF ratio** | **95% CI** |
| --- | --- | --- | --- | --- |
| White | 5409 | 6820 | 0.793 | 0.774 – 0.806 |
| Black | 652 | 2242 | 0.290 | 0.266 – 0.314 |
| Asian | 393 | 187 | 2.102 | 1.868 – 2.332 |
| chi-square goodness-of-fit tests: $p = <0.001$ (0.000) | | | | |
| men | 6584 | 8128 | 0.810 | 0.798 – 0.822 |
| women | 2743 | 1214 | 2.259 | 2.165 – 2.335 |
| chi-square goodness-of-fit tests: $p = <0.001$ (0.000) | | | | |

**Table 3b.** Cardiac Amyloidosis Enrollment Factor Ratio After 2017.

| **Group** | **Observed** | **Expected** | **EF ratio** | **95% CI** |
| --- | --- | --- | --- | --- |
| White | 1339 | 1526 | 0.877 | 0.842 – 0.898 |
| Black | 63 | 502 | 0.125 | 0.108 – 0.152 |
| Asian | 41 | 42 | 0.976 | 0.878 – 1.082 |
| chi-square goodness-of-fit tests: $p < 0.001$ (0.000) | | | | |
| men | 1743 | 1818 | 0.958 | 0.939 – 0.981 |
| women | 301 | 272 | 1.107 | 1.058 – 1.162 |
| chi-square goodness-of-fit tests: $p = 0.013$ | | | | |

**Table 4a.** Phases 0,1, and 2: Cardiac Amyloidosis Enrollment Factor Ratio.

| **Group** | **Observed** | **Expected** | **EF ratio** | **95% CI** |
| --- | --- | --- | --- | --- |
| White | 4233 | 6113 | 0.692 | 0.668 – 0.712 |
| Black | 353 | 2010 | 0.176 | 0.152 – 0.208 |
| Asian | 323 | 167 | 1.934 | 1.614 – 2.246 |
| chi-square goodness-of-fit tests: $p = <0.001$ (0.000) | | | | |
| men | 5618 | 7285 | 0.771 | 0.753 – 0.787 |
| women | 2700 | 1089 | 2.479 | 2.357 – 2.603 |
| chi-square goodness-of-fit tests: $p = <0.001$ (0.000) | | | | |

**Table 4b.** Phase 3 and 4: Cardiac Amyloidosis Enrollment Factor Ratio.

| **Group** | **Observed** | **Expected** | **EF ratio** | **95% CI** |
| --- | --- | --- | --- | --- |
| White | 2515 | 2232 | 1.127 | 1.106 – 1.134 |
| Black | 362 | 734 | 0.493 | 0.453 – 0.527 |
| Asian | 111 | 61 | 1.820 | 1.561 – 2.079 |
| chi-square goodness-of-fit tests: $p = 0.000$ (0.000) | | | | |
| men | 2709 | 2660 | 1.018 | 0.990 – 1.010 |
| women | 344 | 398 | 0.864 | 0.790 – 0.930 |
| chi-square goodness-of-fit tests: $p = 0.004$ | | | | |

Prior to 2017, the EF was as follows: White 0.79 (95% CI 0.77 – 0.81), African American 0.29 (95% CI 0.27 – 0.31), Asian 2.1 (95% CI 1.87 – 2.33), (P value <0.001) and Men 0.81 (95% CI 0.79 – 0.82) and Women 2.25 (95% CI 2.17 – 2.34), (P value <0.001). After 2017 the EF was as follows: White 0.87 (95% CI 0.84 – 0.90), African American 0.13 (95% CI 0.11 – 0.15), Asian 0.98 (95% CI 0.88 – 1.08), (P value <0.001) and Men 0.96 (95% CI 0.94 – 0.98) and Women 1.11 (95% CI 1.06 – 1.16), (P value 0.013).

For phase 3 and 4 clinical trial EF ratios were found to be White 1.12 (95% CI 1.11 – 1.13), African American 0.49 (95% CI 0.45 – 0.53), Asian 1.82 (95% CI 1.56 – 2.08), (P value <0.001), Men 1.0 (95% CI 0.99 – 1.01) and Women 0.86 (95% CI 0.79 – 0.93), (P value of 0.004).

For other phases (0, 1, 2), the EF ratio was found to be White 0.69 (95% CI 0.67 – 0.71), African American 0.18 (95% CI 0.15 – 0.21), Asian 1.93 (95% CI 1.61 – 2.25), (P value <0.001) and Men 0.77 (95% CI 0.75 – 0.79) and Women 2.48 (95% CI 2.36 – 2.60), (P value <0.001).

## Discussion

Our systematic review of clinical trials in ATTR-CM demonstrates persistent and substantial differences in racial and ethnic representation. White and Asian participants were overrepresented, with EFs of 0.81 and 1.91, respectively where EF ≥ 0.75 denotes sufficient representation. By contrast, Black or African American participants exhibited an EF of just 0.26, which represents a 3.11-fold and 7.35-fold lower inclusion than White and Asian participants, respectively. Among sex categories, women were notably overrepresented (EF = 2.05), while men demonstrated adequate inclusion (EF = 0.84). Although representation modestly improved in phase 3 trials (29.4% of studies), where Black participants achieved an EF of 0.49, this still fell short of adequacy and highlighted even greater disparities in earlier-phase trials (EF = 0.12).

Despite significant therapeutic breakthroughs and expanded awareness of ATTR-CM, Black individuals, who account for nearly one quarter of the US ATTR-CM population, remain markedly underrepresented in research participation. In contrast, White and Asian participants are consistently overrepresented, while women are modestly overrepresented relative to disease prevalence. Although the 2017 FDA Final Rule improved demographic reporting transparency, it did not translate into improved minority participation. These findings highlight a widening gap between disease burden and research representation, threatening both the external validity of clinical trial findings and the equitable distribution of therapeutic advances.

Over the past decade, therapies such as tafamidis and acoramidis have transformed ATTR-CM a uniformly fatal condition, to one in which disease modification and prolonged survival are achievable. However, the clinical benefit of these therapeutic breakthroughs cannot be generalized to populations due to insufficient representation in the pivotal studies that established their efficacy. For example, in the pivotal ATTR-ACT and ATTRibute-CM trials evaluating tafamidis and acoramidis, Black participants accounted for only 14.4% and 4.8% of enrollees, respectively [5,12]. These figures fall far short of their estimated disease prevalence of approximately 24% found in the CARS registry [10]. Such imbalances risk limit therapeutic applicability and may perpetuate disparities in outcomes for groups already bearing disproportionate disease burden.

The persistent underrepresentation of Black patients in ATTR-CM clinical trials reflects a complex interplay of inherited predisposition, social determinants of health, and structural inequities. Genetic epidemiology demonstrates a high prevalence of the pathogenic V142I (formerly V122I) transthyretin variant among individuals of West African, Afro-Caribbean, and Caribbean-Hispanic ancestry, contrasting sharply with the predominance of the wild-type variant in White populations [13]. It has also been strongly associated with higher risks of developing heart failure within individuals of Hispanic and African ancestry compared to those without the allele [14]. It’s been further shown that Black patients, particularly those carrying the V142I mutation, face significantly worse event-free survival and elevated incidences of left ventricular assist device implantation, cardiac transplantation, heart failure hospitalization, and all-cause mortality, even after adjustment for disease stage and socioeconomic status (SES) [15]. Socioeconomic disadvantage further compounds this vulnerability. Black patients are disproportionately concentrated in the lowest SES quintiles, and those in high-deprivation neighborhoods experience higher rates of heart failure hospitalization and mortality compared to White patients in similarly disadvantaged contexts [16].

These patterns illustrate that genetic risk is amplified by social inequities, reinforcing the need for early identification strategies among at-risk groups. Large organizations like the American Heart Association have recommended early identification of amyloidosis within at-risk populations such as individuals with Heart failure with preserved ejection fraction (of which majority compromise Hispanic and Black individuals), individuals of West African origin, or those with small fiber polyneuropathy [17].

Even after effective therapies have been developed, access inequities persist. For instance, tafamidis, the first FDA-approved treatment for ATTR-CM carries an annual list price of approximately $225,000 [20]. This cost is nearly four times the median annual household income for Black families ($59,363), compared to White ($93,948) and Asian ($112,800) households. Such cost differentials risk exacerbating existing health inequities, despite proven clinical benefit.

Although the 2017 Final Rule increased demographic reporting from 60% to 85.7%, minority participation paradoxically declined (EF for Black participants decreased from 0.29 to 0.12 post-regulation). In many ways, it failed to confront the deeper structural drivers of racial and socioeconomic underrepresentation in research participation [18,19]. Meanwhile, White and Asian participants consistently met or exceeded adequacy thresholds of EF > 0.75. This disconnect underscores that regulatory transparency alone cannot overcome entrenched barriers rooted in medical mistrust, socioeconomic constraints, and inequitable research infrastructure. Historical ethical violations, such as the Tuskegee Syphilis Study, continues to erode trust in medical research among minority communities. Concurrently, logistical barriers like transportation, financial strain, and limited specialty access, further restrict trial accessibility and participation. Language and cultural discordance between research staff and underrepresented populations further inhibit effective recruitment, particularly when site selection favors predominantly White, affluent regions through convenience sampling. Conversely, the relatively high EFs among Asian participants and women may, in part, reflect sampling biases or registry-based misclassification or a combination of the two causing misrepresentations leading to incorrectly elevated EF values. Asian and women could also be poorly represented within the geographic areas that the tertiary centers contributing to the CARS registry are located.

Addressing these inequities requires a paradigm shift from passive regulatory compliance to proactive, community-engaged strategies. Regulatory reform must be paired with strategies that cultivate and trust and accessibility: community partnerships, engaging with local organizations and minority-serving institutions, culturally tailored education, and practical support such as transportation assistance and flexible scheduling. Research teams should reflect the linguistic and cultural diversity of target populations, all of which are critical for cultivating trust and enhancing trial accessibility among historically marginalized populations [21]. Funding agencies should tie trial approval or continuation to measurable diversity benchmarks. The stakes are high: in an era increasingly governed by value-based care, the continued exclusion of racially and socioeconomically diverse populations contributes to diagnostic delays, preventable hospitalizations, and suboptimal application of therapies, thereby worsening health disparities and inflating long-term healthcare costs. Given that Black participants comprise a significant proportion of the U.S. ATTR-CM population and are more likely to present with advanced disease, ensuring their equitable inclusion in clinical trials is not only a moral and ethical obligation, but a strategic necessity for improving population health outcomes and advancing health system efficiency.

In sum, the current landscape of ATTR-CM research reflects deep-rooted inequities in racial and socioeconomic inclusion. Achieving representative participation demands systemic reform, institutional accountability, and sustained community engagement. By embedding expanded access into the structure of clinical research, the scientific community can ensure that emerging therapies are reflective of and available to the populations they are designed to serve. Achieving representative inclusion is essential not only for scientific validity and therapeutic efficacy, but also for advancing health equity in the management of cardiac amyloidosis.

## Limitations

Several limitations should be acknowledged. Approximately 27.4% of the trials did not report racial or ethnic demographic information potentially biasing the interpretation of enrollment. In addition, our reference point for the observed prevalence of ATTR CM was obtained using data from a single registry 1997 to 2023 however, this is only a limited time frame to draw conclusions for proper epidemiological analysis. This database is comprised of data from only 20 tertiary, mostly academic centers which can exclude a percentage of patients with cardiac amyloidosis especially from the community-based practices. This can affect the generalizability of our analysis given we were unable to compare our trials to true population level prevalence of the disease within various demographics, like in the case of women and Asians.

Additionally, our study relied solely on publicly available data extracted from clinicaltrials.gov. Race is typically self-reported in trials, which may introduce some misclassification or lack of classification. This is especially true with the Latino population, which is a meaningful subgroup with a known ATTR genetic predisposition, accounted for 1.9% of reported patients in our study but given lack of CARS data on this population we were unable to calculate an EF for this group. Future studies should integrate multi-database approaches and require standardized, transparent demographic reporting to enable comprehensive equity assessment.

Although only US based trials were utilized, some trials were conducted in multiple countries (including the US) which could have caused some misrepresentation with the variation in demographic representation in the countries included, for example a higher proportion of white individuals would be expected in European countries as compared to the US.

## Conclusion

This study highlights profound and persistent imbalances in racial and ethnic representation within US cardiac amyloidosis clinical trials, particularly the under-enrollment of Black participants. Although the 2017 FDA Final Rule improved demographic reporting, it did not improve enrollment of minorities. Although later phase trials demonstrated modest progress, no phase achieved adequate representation. Ensuring equitable inclusion requires a comprehensive strategy that combines community engagement with targeted recruitment strategies and policy adjustments. Only through deliberate structural change can clinical trials generate evidence that is both scientifically valid and socially just.

## Data Availability

The data used in this manuscript can be found in the publicly available clinicaltrials.gov

## Statements and Declarations

The authors have no financial disclosures.

## Figures and Tables

### Prisma figure showing project outline

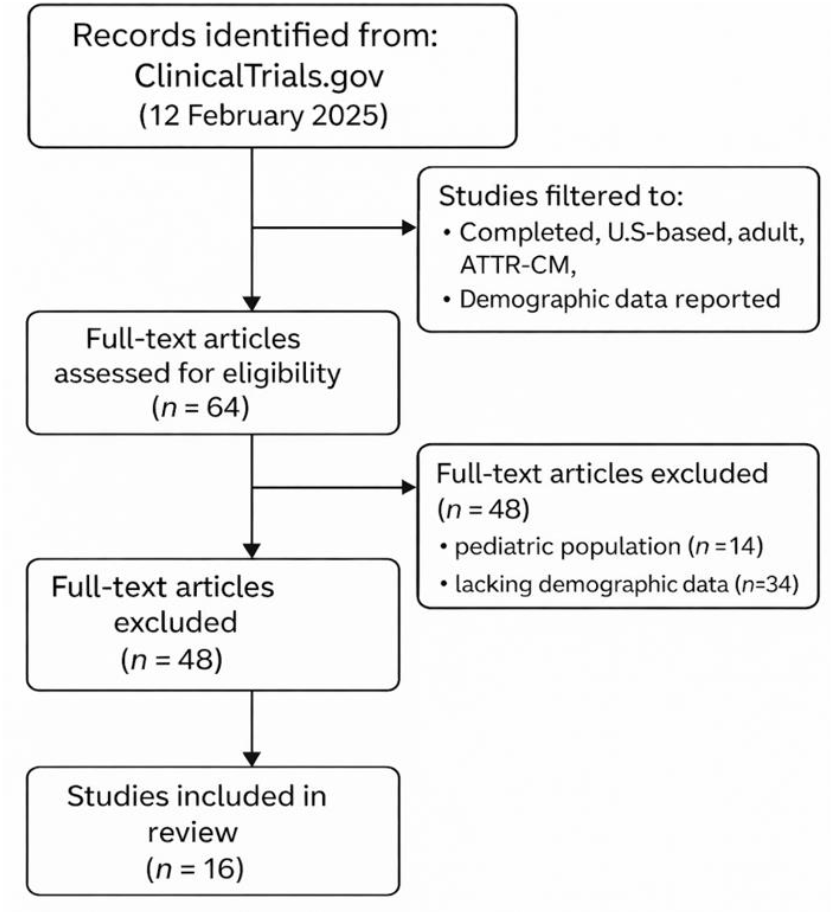

